# Safety, Immunologic Effects and Clinical Response in a Phase I Trial of Umbilical Cord Mesenchymal Stromal Cells in Patients with Treatment Refractory Systemic Lupus Erythematosus

**DOI:** 10.1101/2022.03.31.22273106

**Authors:** Diane Kamen, Caroline Wallace, Zihai Li, Megan Wyatt, Crystal Paulos, Chungwen Wei, Hongjun Wang, Inaki Sanz, Bethany J Wolf, Paul J Nietert, Angela Robinson, Gary Gilkeson

## Abstract

**Background:** Reports of clinical improvement following mesenchymal stromal cell (MSC) infusions in refractory lupus patients at a single center in China led us to perform an explorative Phase I trial of umbilical cord derived MSCs in patients refractory to six months of immunosuppressive therapy.

**Methods:** Six women with a SLEDAI>6, having failed standard of care therapy, received one IV infusion of 1×10^6^ MSCs/kg of body weight. They maintained their current immunosuppressives, but their physician was allowed to adjust corticosteroids initially for symptom management. The clinical endpoint was an SRI of 4 with no new BILAG As and no increase in Physician Global Assessment score of >0.3 with tapering of prednisone to 10mg or less by 20 weeks.

**Results:** Of 6 patients, 5 (83.3%; 95% CI = 35.9% to 99.6%) achieved the clinical endpoint of an SRI of 4. Adverse events were minimal. Mechanistic studies revealed significant reductions in CD27IgD negative B cells, switched memory B cells and activated naïve B cells with increased transitional B cells in the 5 patients who met the endpoint. There was a trend towards decreased autoantibody levels in specific patients. One patient had an increase in their Helios+Treg cells, but no other significant T cell changes were noted. GARP-TGFβ complexes were significantly increased following the MSC infusions. The B cell changes and the GARP-TGFβ increase were significantly correlated with SLEDAI scores.

**Conclusion:** This pilot trial suggests that UC MSC infusions are safe and may have efficacy in lupus. The B cell and GARP-TGFβ changes provide insight into mechanisms by which MSCs may impact disease.

**Trial Registration:** NCT03171194

**Funding:** This study was funded by a grant from the Lupus Foundation of America and NIH UL1 RR029882

## Introduction

Systemic lupus erythematosus (SLE) is a heterogeneous disease affecting young women in their childbearing years (1). The hallmark of disease is production of autoantibodies with immune complex deposition in target organs. Despite research progress and recent clinical trials’ success, there is still a need for effective safe treatments (2, 3). Current immunosuppressive and biologic therapies have therapeutic effects, yet a significant number of lupus patients remain inadequately responsive to current therapies. An additional issue with current therapies is the side effect profile especially for women of childbearing potential (4). Cellular therapies, such as mesenchymal stromal cells (MSCs) are an emerging area of interest as to their therapeutic efficacy in immune diseases including lupus.

MSCs are derived from bone marrow, adipose tissue and umbilical cords/placentas (5-7). Their local autologous use in plastic and orthopedic surgery is proven beneficial (8, 9). There is growing literature on the immune properties of MSCs and their use in immune-mediated diseases (10, 11). Trials of MSCs in refractory graft versus host disease, rheumatoid arthritis, inflammatory bowel disease and lupus have had variable results (12-17). Most were uncontrolled trials with small numbers of participants. There is a benefit of MSCs for steroid refractory pediatric graft versus host disease (18) and local use in healing fistulas in Crohn’s (19). The efficacy in other diseases remains unproven due to a lack of placebo-controlled trials.

There are a number of publications regarding use of MSCs in refractory lupus from a single center in Nanjing, China (15, 20, 21). The reports provide both short and long-term follow-up of dozens of patients treated with MSCs. There was an overall response rate of 60-65% at six months following a single MSC infusion of one million cells per kilogram. Long-term beneficial effects on disease activity were reported (15, 21, 22). The patients primarily had lupus nephritis, but other manifestations of lupus were also improved. The length of response varied from 6 months to five years (23). None were placebo controlled. The one controlled trial of MSCs in lupus nephritis, enrolled treatment naïve patients, was small (18 patients) and based on a high response rate to cyclophosphamide alone versus cyclophosphamide plus MSCs did not detect an added benefit of MSCs(24).

MSCs advantages are they are easily obtained, have a low side effect profile and can be given without histocompatibility matching or pre-conditioning (25). The reported “immune privilege” of MSCs is based on their not expressing MHC Class II or immune cofactors, rendering them initially hidden from the host immune system(26). There are literally dozens of proposed mechanisms for the immune effects of MSCs, though none are proven in humans (27, 28).

Due to the promising results out of China, we initiated studies of MSCs in lupus, starting with murine models, that demonstrated efficacy of MSCs from human controls in reducing renal disease(29). Allogeneic MSCs were used in this Phase I trial as autologous MSCs from lupus patients are not as immune active as allogeneic MSCs (30). In a limited study of autologous bone marrow MSCs, two lupus patients did not have a beneficial effect on their disease (31). Based on the lack of definitive evidence of MSC efficacy, we performed a Phase I safety trial in treatment refractory lupus as a preliminary assessment in a multi-ethnic cohort.

## Methods

### Preparation of UC MSCs

The MSCs were derived from UCs of two healthy donors under FDA IND 16377. The donors were mothers in the OB/GYN clinic undergoing elective C-sections. After informed consent, the mother’s blood was tested using the infectious testing battery required for allogeneic bone marrow donors within one week of delivery. Antinuclear antibody (ANA) screening was done. Potential donors were excluded if they had personal or family history of autoimmune disease or any positives on infectious and autoimmune testing of their blood. The cords came from one male and one female infant. The UCs were obtained using sterile technique and transported to the MUSC Center for Cellular Therapy (CCT). The derived cord cells were plated and incubated in a minimal essential medium (MEM, GIBCO) with glutamine and 10% sterile pooled human platelet lysate (Cook Regenec Inc). Aliquots were tested for bacterial, fungal, endotoxin, and mycoplasma. MSC immune potency was measured by T cell proliferation and interferon-gamma induced IDO expression. Further details are provided in the Supplemental Methods.

### Patients

Patients had a historical presence of at least 4 of 11 of the ACR Lupus Classification Criteria **(32)**. Further inclusion criteria included: age between 18 and 65 years old, either sex, any race, evidence of a positive ANA (≥1:80 titer) or positive dsDNA antibody test within 6 months of screening, clinically active SLE determined by SLEDAI score ≥6 and ≤12, and the presence of one BILAG A or one BILAG B at screening, despite standard of care (SOC) therapy. If the BILAG A or the BILAG B was in the renal organ system, the patient must have completed 6 months with either mycophenolate mofetil or cyclophosphamide. Non-nephritis patients had active disease despite 3 months of SOC therapy. Patients were able and willing to give written consent. Details regarding patient selection are in the Supplemental Methods and in Table 1. A sample size of n=6 was selected in an attempt to balance the need to investigate the safety of this therapy with the need to limit any negative consequences should they occur.

**Table 1:** Baseline demographics and disease characteristics in the six participants. Demographics of the participants is presented in Column 2 with baseline lupus manifestations and disease duration are shown in Columns 2 and 3. There was a wide range of disease manifestations and disease duration. Baseline medications, SLEDAI at baseline and SLEDAI at week 24 are presented in Columns 5, 6 and 7

| Subject | Non-Serious AEs<br>(# NCI-CTCAE Grade) | SAE (#) | AEs Related to MSCs | Early Withdrawal |
| --- | --- | --- | --- | --- |
| 1 | 3 Grade 1<br>5 Grade 2<br>0 Grade ≥3 | 0 | Grade 2 Nausea<br>("possible") |  |
| 2 | 0 Grade 1<br>2 Grade 2<br>0 Grade ≥3 | 0 |  |  |
| 3 | 2 Grade 1<br>2 Grade 2<br>0 Grade ≥3 | 1 | Grade 1 Paresthesias<br>("possible")<br>Grade 1 Flushing<br>("possible") | Dropped out after Week 8 visit |
| 4 | 0 Grade 1<br>3 Grade 2<br>0 Grade ≥3 | 0 | Grade 2 Tachycardia<br>("possible") |  |
| 5 | 1 Grade 1<br>1 Grade 2<br>0 Grade ≥3 | 0 |  |  |
| 6 | 1 Grade 1<br>1 Grade 2<br>0 Grade ≥3 | 0 |  |  |
| TOTAL | 21 AEs | 1 SAE | 4 AEs possibly treatment<br>related; all resolved<br>without sequela | 1 early withdrawal to pursue other SLE<br>treatments at Week 8 |

## Clinical endpoints

### Clinical Response

The (SLE Responder Index) SRI 4 was used as the assessment tool for clinical activity. A decrease in the SLEDAI of at least 4, no new BILAG As or two BILAG Bs and no increase in the Physicians Global Assessment >0.3 were required to be considered responsive. This assessment was made at weeks 0, 4, 8 and 24 after the MSC infusion. The week 24 assessment was the primary endpoint. Inability to taper prednisone to 10mg or less by 20 weeks was considered a treatment failure. Dose increases or new additions to SOC immunosuppressant therapy for SLE activity prior to Week 24 were considered a treatment failure. Secondary outcomes included the SF-36 quality of life instrument and the Lupus Impact Tracker(33).

### Safety

Study participants reported adverse events (AEs) throughout the trial, regardless of attribution. Lowering of standard of care (SOC) immunosuppressant therapy due to toxicity was allowed. Further safety methodology is in the Supplemental Methods.

### Treatment protocol

All patients received UC-derived MSCs suspended at a concentration of 2 × 10^6^ cells/mL in Plasma-Lyte A (Baxter) suspension media. The patients and the treatment team were aware they were all receiving MSCs. The patients received 1 × 10^6^ cells/kg body weight. The infusion rate was 100 × 10^6^ over 10 minutes. Patients received premedication of Benadryl 25mg and 650mg of Tylenol orally. There was no preconditioning or HLA matching. If the patient was CMV antibody negative, they received cells from the donor that was CMV negative. If the patient was CMV antibody positive, but not having an acute infection, they received cells from the CMV positive donor. Further description of the treatment protocol is in the Supplemental Methods.

### Statistical analysis

The primary endpoint consisted of the proportion of participants who exhibited a clinical response at week 24 by SRI4. This proportion was reported along with an exact 95% confidence interval. All secondary analyses were conducted in an exploratory fashion with p-values and confidence intervals presented without adjustments for multiple comparisons. Interval estimates were generated at the 95% confidence level.

Since many of the secondary endpoints were collected at multiple time points, statistical models appropriate for longitudinal data analyses were used (34). General linear mixed models (GLMMs), included appropriate covariance structures to account for within-subject clustering, were constructed for the different outcomes to determine whether there were significant changes over time (i.e., for the SLEDAI, SF-36, LIT) and whether certain outcomes were correlated with others (i.e., B/T cell subtype distributions and autoantibody levels). Sensitivity analyses were conducted by adopting a last observation carried forward (LOCF) approach within the GLMM models, given that one subject (#3) did not contribute data after week 8.

### Mechanistic Studies

Protocols for handling of specimens, B cell and T cell characterizations, ELISA assays and Glycoprotein A repetition predominant (GARP) assays are in the Supplemental Methods section.

## Results

### Safety

As shown in Table 1, there were a total of 21 AEs over the 52 weeks of the trial. None were Grade 3 or higher and only 4 were felt possibly related to the MSC infusions and all resolved quickly. These included mild nausea, paresthesias and flushing. There were no lab-related AEs in the six participants. The one non-responder (Patient 3) was treated with Rituximab by her primary rheumatologist four months post-MSC infusion for refractory symptoms and had an anaphylactic reaction. She lived in California and did not want to make the cross-country trips post week 8. This was the only SAE in the trial and was judged not due to MSC treatment given a prior history of multiple anaphylactic reactions to intravenous medications. There were no other common AEs within the group.

### Clinical Response

The six patients enrolled were female with an average age of 38 (range of 26 to 48). Two participants were African American, one was Hispanic and three were Caucasian. Average disease duration was 8.2 years (range of 3.9 to 11.7 years). One patient had onset of disease as a child. Baseline disease features are in Table 2. One patient had refractory episodes of transverse myelitis despite immunosuppression and biologic therapy. One patient had renal disease with ongoing proteinuria post-therapy with mycophenylate (MMF). All but one patient were on hydroxychloroquine and prednisone. Two were on MMF, one on azathioprine and MMF and one on cyclosporine. Two were not on an immunosuppressant having failed multiple immunosuppressive regimens. All patients continued their baseline medications throughout the trial.

**Table 2:**
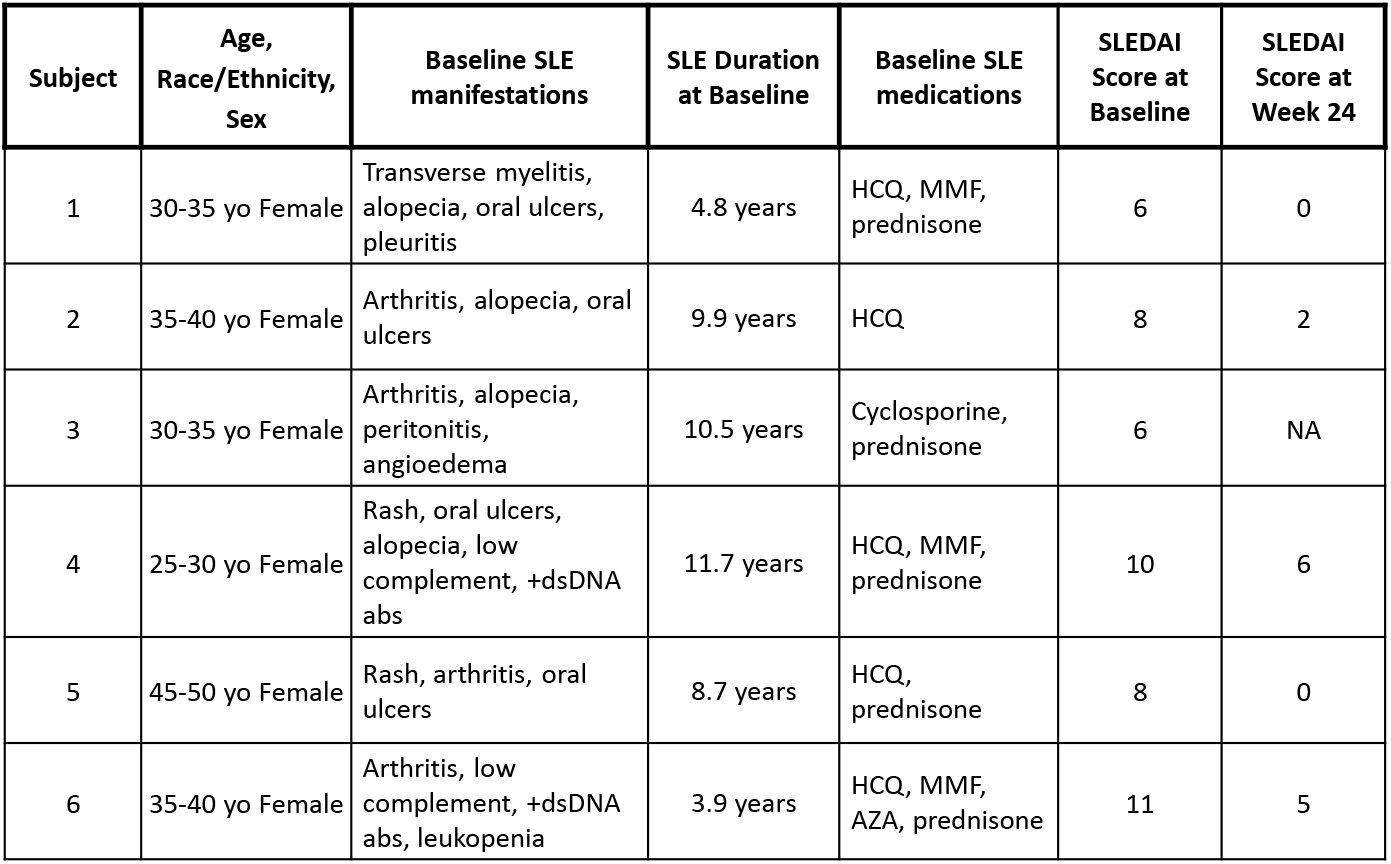
Change in the physician global assessment and prednisone dosing. The change in the PGA from baseline to week 24 is presented in Column 2, while the change in prednisone dosing from baseline to week 24 and to week 52 is presented in Columns 3 and 4. Three of the five responders were able to taper prednisone to 5mg or less while two maintained their prednisone dose at 10mg/day.

ble 2: Baseline Demographics & Disease Characteristics
| Subject | Age,<br>Race/Ethnicity,<br>Sex | Baseline SLE<br>manifestations | SLE Duration<br>at Baseline | Baseline SLE<br>medications | SLEDAI<br>Score at<br>Baseline | SLEDAI<br>Score at<br>Week 24 |
| --- | --- | --- | --- | --- | --- | --- |
| 1 | 30-35 yo Female | Transverse myelitis,<br>alopecia, oral ulcers,<br>pleuritis | 4.8 years | HCQ, MMF,<br>prednisone | 6 | 0 |
| 2 | 35-40 yo Female | Arthritis, alopecia, oral<br>ulcers | 9.9 years | HCQ | 8 | 2 |
| 3 | 30-35 yo Female | Arthritis, alopecia,<br>peritonitis,<br>angioedema | 10.5 years | Cyclosporine,<br>prednisone | 6 | NA |
| 4 | 25-30 yo Female | Rash, oral ulcers,<br>alopecia, low<br>complement, +dsDNA<br>abs | 11.7 years | HCQ, MMF,<br>prednisone | 10 | 6 |
| 5 | 45-50 yo Female | Rash, arthritis, oral<br>ulcers | 8.7 years | HCQ,<br>prednisone | 8 | 0 |
| 6 | 35-40 yo Female | Arthritis, low<br>complement, +dsDNA<br>abs, leukopenia | 3.9 years | HCQ, MMF,<br>AZA, prednisone | 11 | 5 |

Overall, 5 (83.3%; 95% CI = 35.9% to 99.6%) of the 6 participants receiving UC MSCs reached the primary response criteria of an SRI of 4 by 24 weeks and a decrease in prednisone to 10mg a day or less by 20 weeks (Figure 1A, Table 3 and Supp Table 1). By week 24, results from the GLMMs showed that there was a significant (p<0.001) decline from baseline in the SLEDAI scores, decreasing from a baseline average of 8.2 (range 6 to 11) to 2.8 (range 0 to 6) at week 24, for a mean decline of 5.3 units (95% CI = 2.7 to 8.0). Significant (p<0.05) and sustained responses in the SF-36 scores and the Lupus Impact Tracker over time were observed (Figures 1B and 1C). Sensitivity analyses using the last observation carried forward approach for missing data yielded results that were similar to the primary analyses; significance (p<0.05) was preserved for the time changes noted in the SLEDAI; the SF-36 general health, social functioning, and vitality domains; and the Lupus Impact Tracker.

**Table 3:** Safety reports during the trial including AEs and the one SAEs. There were no Grade 3 or higher AEs. AEs deemed definitely not related to the investigational product are totaled numerically but not detailed in the table. Attribution of the AEs and SAE is presented in Column 3

| Subject | Physician's Global Assessment<br>Change over 24 Weeks<br>(PGA, scale 0-3) | Baseline<br>Prednisone Dose<br>(mg/day) | Week 24<br>Prednisone Dose<br>(mg/day) | Week 52 Prednisone<br>Dose (mg/day) |
| --- | --- | --- | --- | --- |
| 1 | **-1.94** | **10** | **7.5** | **5** |
| 2 | **-0.9** | 0 | 0 | 0 |
| 3 | n/a | 20 | -- | -- |
| 4 | **-1.23** | **20** | **10** | **2.5** |
| 5 | **-1.05** | 10 | 10 | 10 |
| 6 | **-1.8** | 10 | 10 | 10 |

**Figure 1.**
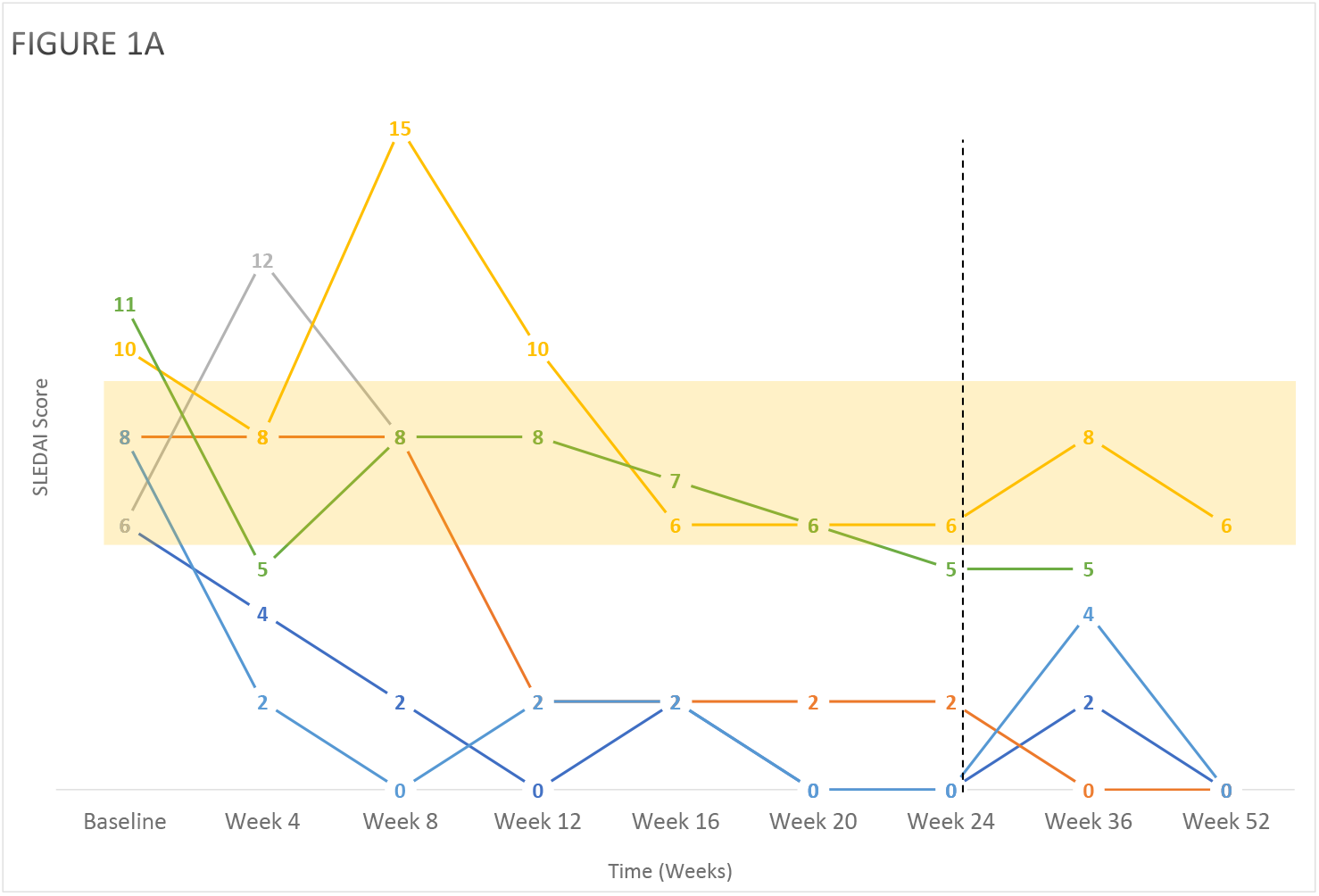
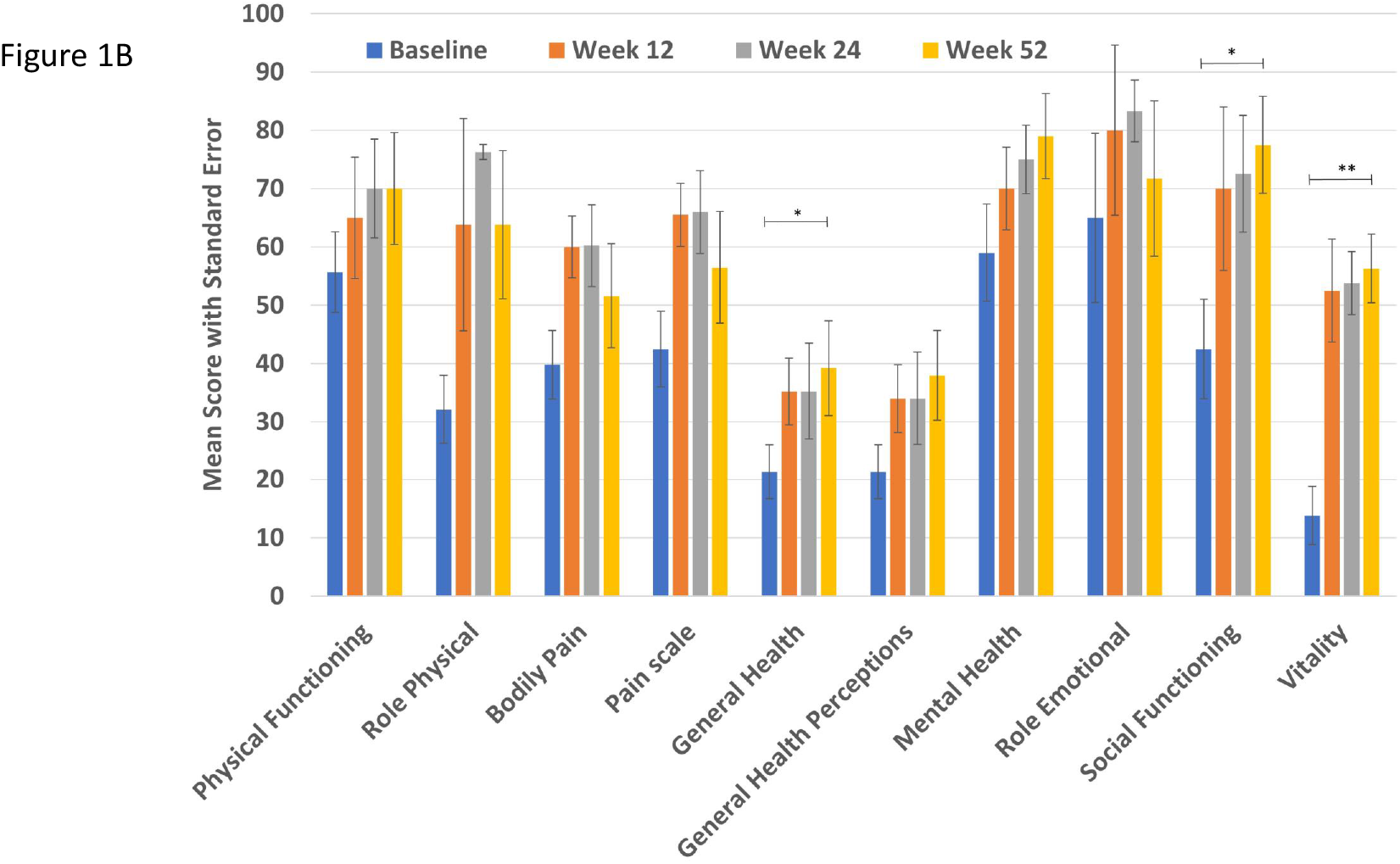
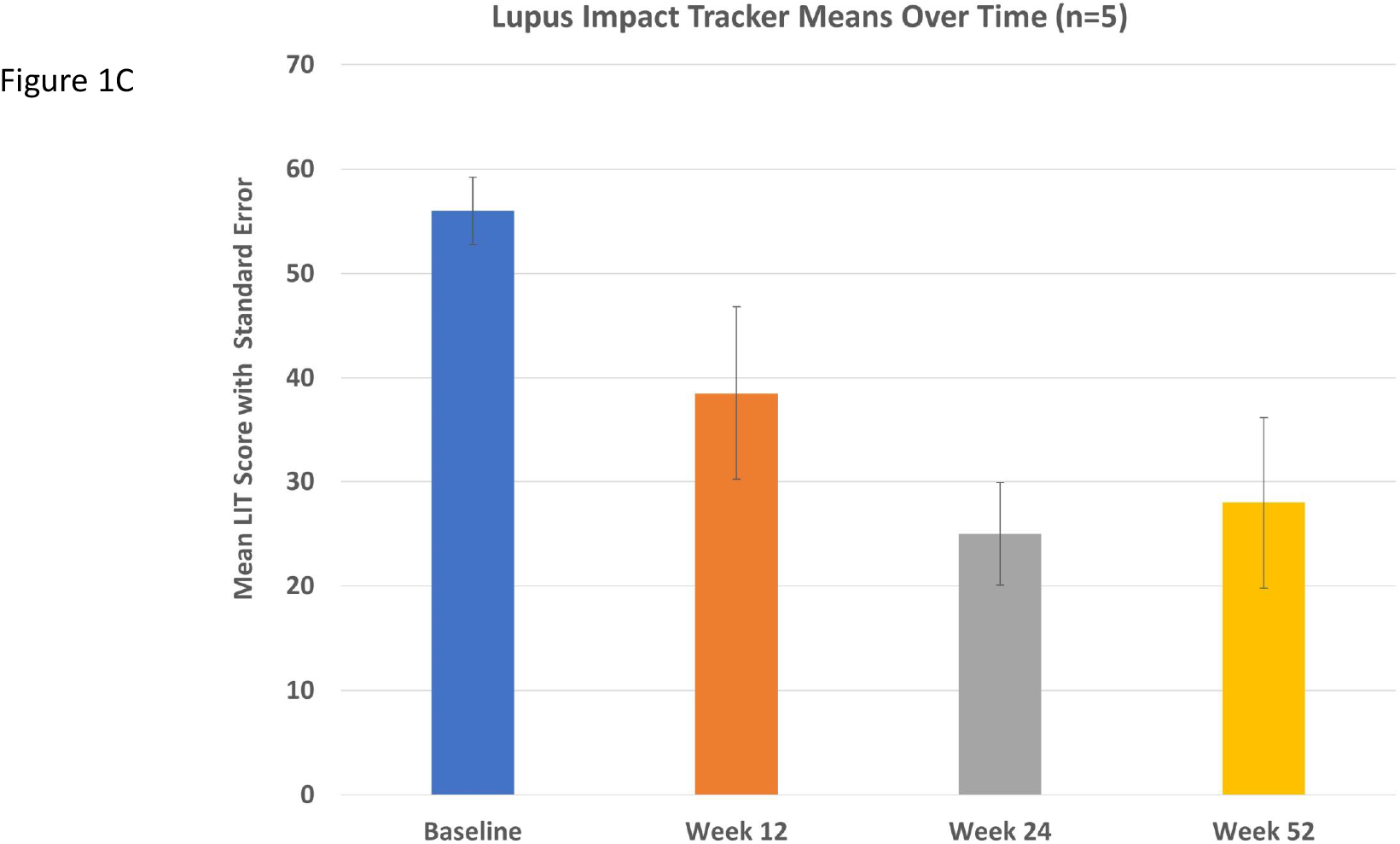
Clinical and Patient Reported Outcomes. **Fig 1A** Line plot of the change in SELENA-SLEDAI scores over the course of the trial. This is the overall score combining lab and clinical criteria. The primary endpoint was the SLE responder index (SRI-4 at 24 weeks). There was overall a significant decline in SELENA SLEDAI scores of 5/6 patients meeting the SRI-4 endpoint (p<0.006). **Fig 1B** Lupus Impact Tracker Means over the time of the study. There was a significant decrease in the LIT beginning at week 12 and continuing through week 52 (p=0.007). Error bars indicate standard deviation. Represents the scores of the 5 patients completing the trial to 52 weeks. **Fig 1C** SF-36 Subscale scores at the given timepoints for the five patients who completed the study. Significant increases in general health (p=0.02), social functioning p=0.02) and vitality (p=0.004) were present beginning at week 12 and continuing through week 52. Data represents the scores of the 5 patients completing the trial to 52 weeks

Standard laboratory measures were assessed. Participant 3s proteinuria improved from baseline (1015 mg/g/day) to week 24 (192 mg/g/day) with an increase in her lymphocyte count from 290 to 1000 and her C4 from 9.5 mg/dl to 13.1 mg/dl. Participant 6 had a decrease in her anti-dsDNA level from a baseline of >300 IU/ml to 132 IU/ml at week 24.

As shown in Figure 1B, titers for anti-Ro52, anti-Ro60, anti-Sm and anti-RNP were assessed at each of the 0-, 4-, 8- and 24-week timepoints. All the participants had increased titers of anti-Ro52, anti-Ro60 and anti-RNP greater than control. Participants 5 and 6 had titers of anti-Sm elevated above control. In participants 2 and 6, there was a log-fold decrease in anti-Ro60 antibodies between week 0 and week 4 that remained through week 24. Other titers remained stable over the time of the study.

There was stability or further improvement in the clinical response from 24-52 weeks in the 5 responders (Figure 1A and Supp Table 1). Physician global and patient global assessments were significantly improved in the 5/6 responders (Table 3 and data not shown). Prednisone was able to be tapered or maintained at 10mg or less per day (Table 3). There was a sustained response in the Lupus Impact Tracker (Figure 1C). Subsequently, from 18-48 months after completion of the study, 4 of the patients flared. Patient 1 had a recurrence of her thoracic cord transverse myelitis at 20 months post infusion. She was treated with Cytoxan and pheresis and retreated with MSCs. She has had no flares now 34 months later. The other three participants had less severe flares with arthritis and skin disease and were not retreated with MSCs.

### Mechanistic studies

#### B cell responses

Flow cytometry was performed on patient samples at weeks 0, 4, 8 and 24. Week 8 data is not included due to weather induced loss of three week 8 samples in transit. The gating scheme was previously published with the identification of nine B cell subsets (35): 1. Plasmablasts, 2. Double negative 3+4 (DN 3+4), 3. Double negative 2 (DN2), 4. Double negative 1 (DN1), 5. Switched memory (SM), 6. Unswitched memory (USM), 7. Activated naïve (aN), 8. Resting naïve (rN) +transitional 3 (T3), and 9. Transitional 1+ 2 (T1+T2).

As shown in Figure 2, there was variation in percentage of B cell subsets at baseline. A significant change from week 0 to 24 was a marked decrease in the percentage of total DN B cells in participants 1, 2, 5 and 6 (Figure 2 and Supp Table 2). DN2 B cells are expanded in African American women with active lupus (35). Of interest, the two patients with the highest numbers of DN2 cells at baseline were the two African American participants (1 and 6). There was a significant reduction in DN2 B cells following MSC infusion. aN B cells are also increased in African American females with lupus (33). Three of the participants (1, 5, and 6) had detectable numbers of aN B cells. Participants 1 and 6 had expanded DN2 B cells and aN B cells in parallel through the study.

**Figure 2.**
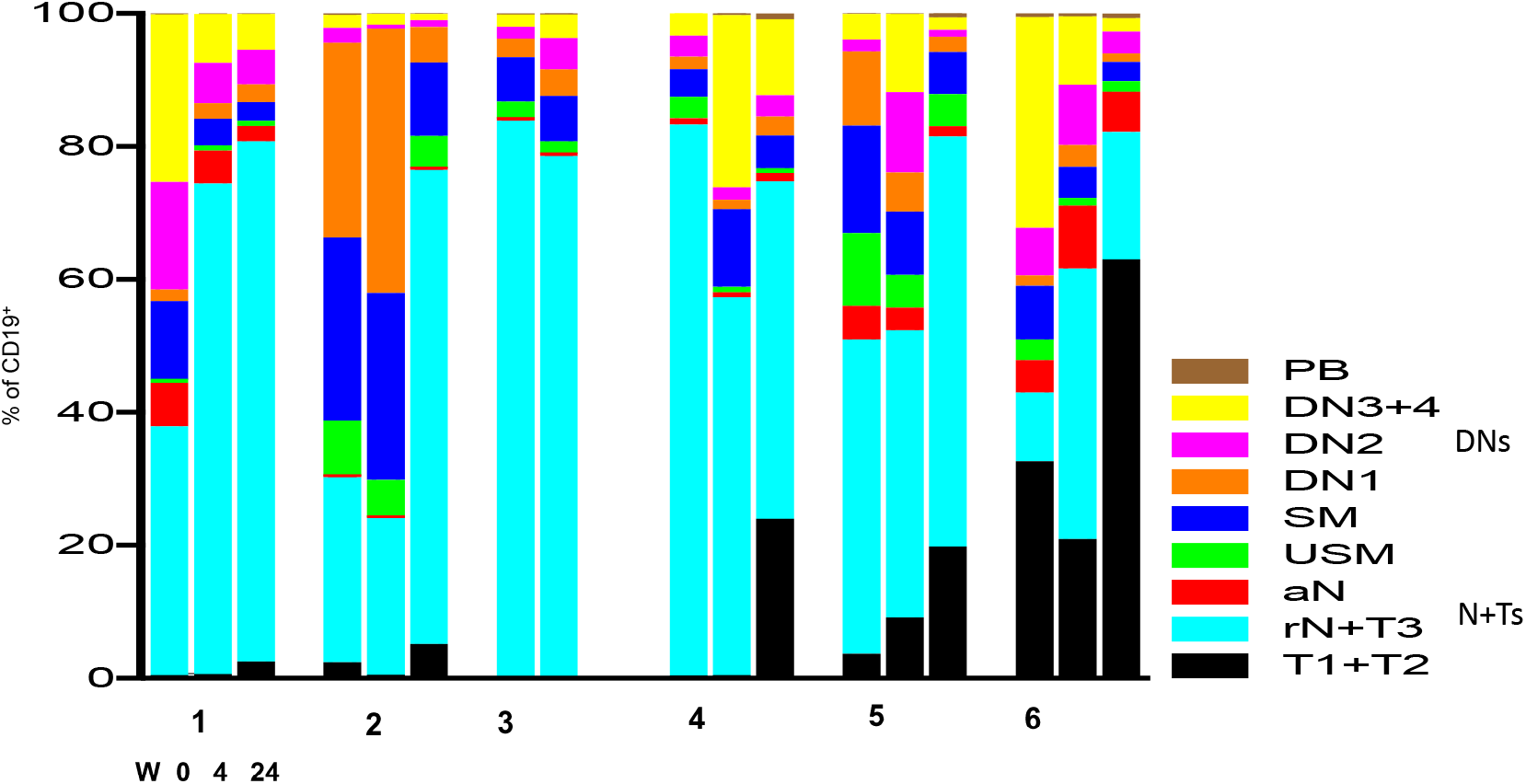
B cell subset changes over time by Flow Cytometry. B cell subset changes over time as determined by flow cytometry. Significant differences in B cell subsets occurred over the trial period as also noted in Table 4. The cell markers used are listed in the methods and are previously published (33). Most significant changes were in decreased double negative B cells including DN2, increased transitional B cells, and decreased activated naive B cells. IgD^-^CD27^+^=SM+PB, IgD^-^CD27^-^=DN1+DN2+(DN3+4), IgD^+^CD27=(T1+T2)+(rN+T3)+aN PB=plasmablasts, DN= double negative, SM= switched memory, USM= unswitched memory aN= activated naïve; rN= resting naïve; T= transitional

Concomitant with the decrease in DN and aN B cells, there was an increase in resting naïve and T1+T2 B cells. There was a significant change in the SM B cells, decreasing in all five of the patients that were responders. Table 5 presents the B cell data in a numerical format including the SLEDAI score calculated at the week 0-, 4-, and 24-week visits. There were significant associations between subjects’ SLEDAI scores and their percentages of N+T (p=0.042), SM (p=0.007) and DN (p=0.041) B cells over time. There was a negative correlation for N+T with the SLEDAI and positive correlation of the SM and DN with the SLEDAI.

### T cell responses

Prior studies of MSC infusions in lupus-prone mice, and in more limited studies in human lupus, reported an increase in Treg cells with a decrease in Th17 and T follicular helper (Tfh) cells following MSC infusions (36).

As shown in Figure 6, we assessed the fold change of Treg cells in the peripheral blood of the participants. Only in patient 1 was there a significant increase in percentage/fold change in Treg cells. This change was present in both Helios- and Helios+ Treg cells. Participant 6 also had an increase in Treg cells present only at week 24 and primarily in Helios-Treg cells. Participant 2 also had a consistent increase in her Tregs as measured by fold change compared to baseline.

Figure 7 demonstrates there were fluctuations in Th1, Th2 and Th17 cells, but no clear trend or significant change during the study. In Figure 7, we also present measures of Tfh and T peripheral helper (Tph) cells. As expected, there were very few Tfh cells detected, although participant 1 had a sustained decrease following MSC infusion. A limited number of Tph cells were detected and there was no significant change. There were no associations between SLEDAI score and changes in T cell subsets other than in Treg levels in patients 1 and 2. There were no detectable changes in CD8+ T cells or their subsets, nor NKT cells (data not shown).

Glycoprotein A repetition predominant (GARP) is a cell surface protein that is a repository for latent TGFβ (LTGFβ) and plays a key regulatory/tolerance role in immunity via modulating TGFβ activation (37). It is primarily expressed on platelets, Tregs, activated B cells and MSCs (38, 39). Lack of GARP expression on murine B cells or Treg cells results in lupus-like autoimmunity (39). We postulated that GARP was involved in the impact of MSCs on the immune response. We assessed the presence of soluble GARP-TGFβ complexes in the serum of lupus patients from our biorepository that were not in the trial. There was a significant (p=0.023) decrease in serum levels of circulating GARP-TGFβ complexes in lupus patients versus controls (Figure 3B, Graph A). We then assessed if there was a correlation between circulating GARP-TGFβ levels and disease activity in these biobank patients. As shown in Figure 3B Graph B, there was a significant inverse correlation (p=0.034) between serum levels of soluble GARP-Latency Associated Peptide (LAP), and SLEDAI scores in patients with active disease (SLEDAIs >10). In Figure 3B Graph C, prior to infusion, serum GARP levels were undetectable in the six participants. At week 4, GARP-TGFβ serum levels were markedly increased from baseline in all patients. At week 8, levels fell in all the patients but remained above baseline. At week 24, there was an upward rebound or stability of GARP levels in 4 of the 5 patients that completed the study (1, 2, 3 and 5).

**Figure 3.**
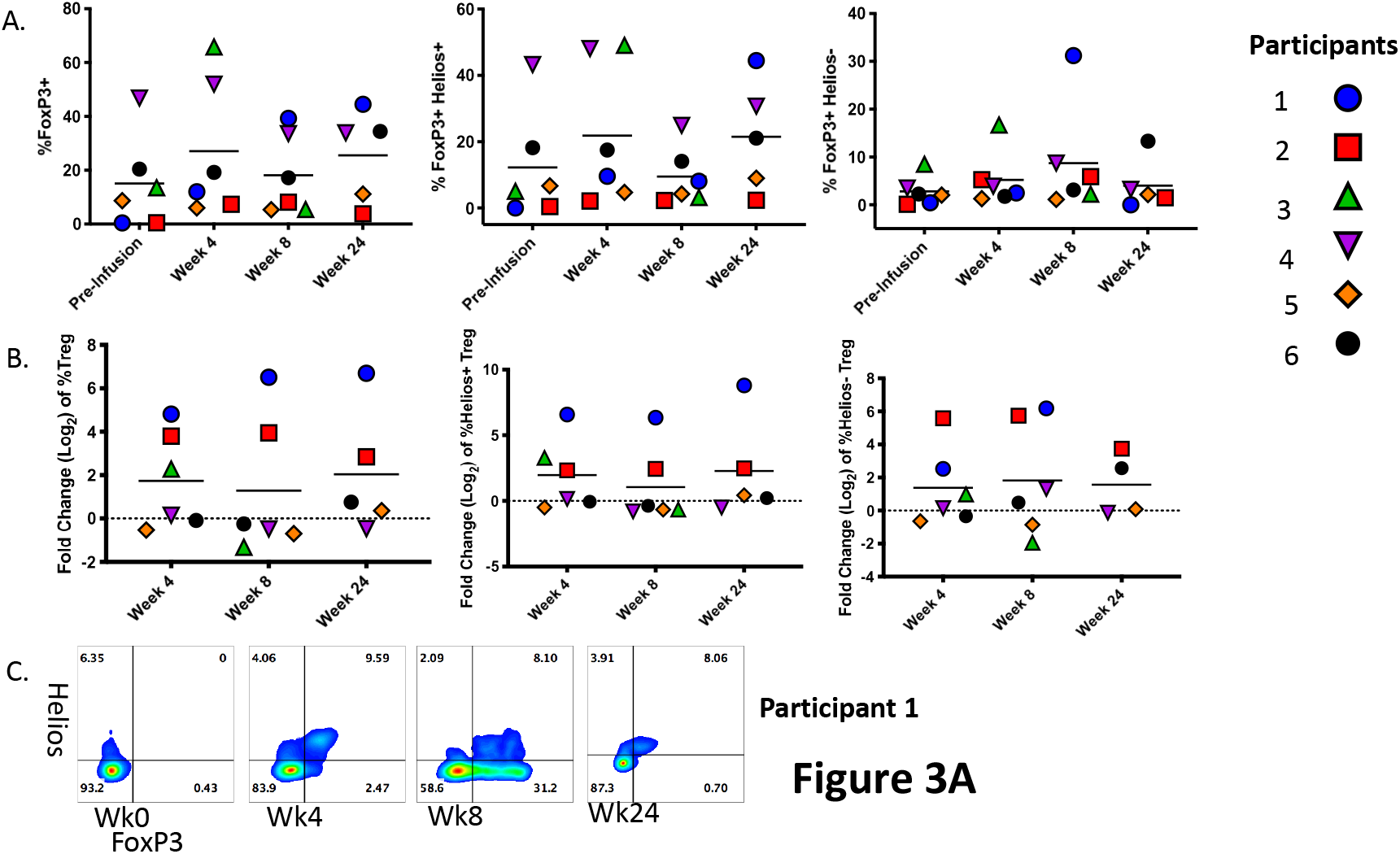
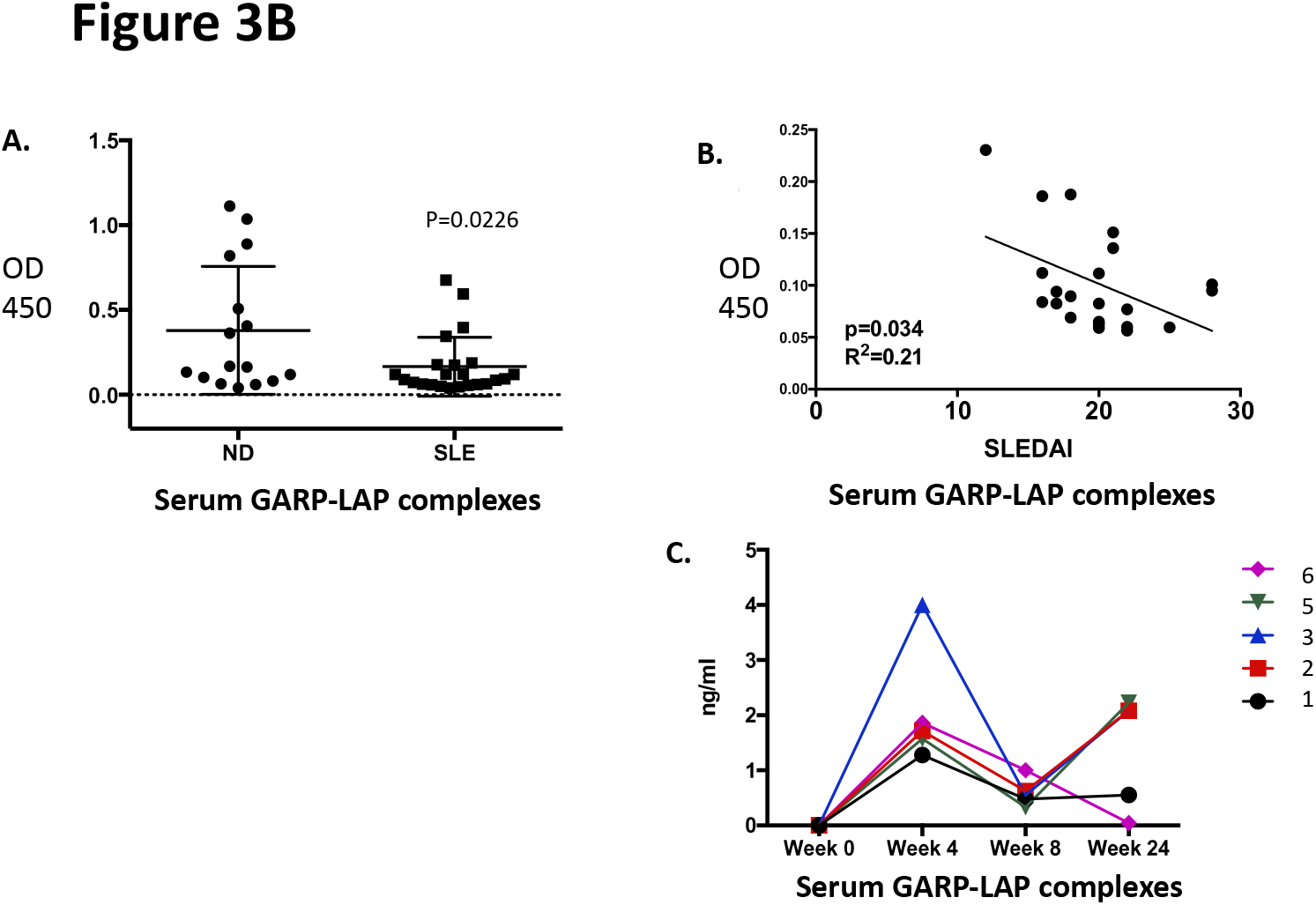
T cell and GARP/TGFb changes over time. **Fig 3A** Treg changes over time are presented. Participant 1 had a significant increase over time in Tregs both Helios+ and Helios-looking at percentage change and fold change over baseline. Participant 2 had a significant increase with time compared to baseline in both Treg subsets, though the percentage change was small given the low percentage of Tregs at baseline. A representative flow block is shown in Figure 7C demonstrating the increase in Tregs over time from 0 to 8% in participant 1 **Fig 3B** GARP serum measures. Panel A shows measures of circulating serum GARP-LAP (latency associated peptide) complexes as measured by a sandwich ELISA assay as previously described (37). Random SLE patients in the MUSC cohort (n=30) had significantly less (p=0.0226) circulating GARP-LAP complex than age/sex/race matched controls (n=16) y axis is OD 450. Panel B plots GARP-LAP complexes via ELISA versus patient SLEDAI score at the time of blood draw (n=21) for the MUSC lupus cohort. The y-axis is OD 450 reading via ELISA. There is a significant correlation between SLEDAI score and GARP-LAP complexes in patients with SLEDAI scores >10. Panel C is a sandwich ELISA measure of soluble serum GARP expressed as ng/ml of serum in the MSC treated patients over time demonstrating near 0 levels of serum GARP at baseline with significant increases at week 4, decreasing at week 8 prior to rebounding at week 24 in 3/5 patients. Mann Whitney U test was used to determine significance (week 0-week4- p=0.003)

## Discussion

This Phase I trial is the first of allogeneic MSCs performed in multi-ethnic lupus patients. The results indicate that infusion of allogeneic UC MSCs appears safe short term, as we had no serious adverse events that were attributed to the UC MSC infusions, and all the AEs were Grade 2 or less. We were encouraged that 5 of the 6 patients treated met the primary endpoint of an SRI 4, justifying performing the Phase II double-blind multi-center efficacy study currently in progress. We were also encouraged by the marked B cell changes and increased serum GARP-TGFβ measures noted suggesting the MSCs had a systemic immune effect.

The patients in this trial were of mixed ethnicity with a range of ages. The patients had variable lupus manifestations. The prior studies in China by Dr. Sun included patients with primarily refractory lupus nephritis, but also included patients with significant hematologic involvement and pulmonary hemorrhage (15, 21, 22). In a recent review of his cohort, Dr. Sun reported that younger patients and those with musculoskeletal symptoms were not as responsive to UC MSC infusion as other lupus manifestations (20).

The duration of response is variable in the reports from Dr. Sun’s group (15, 21, 22). He reported a 65-70% early “response” rate with a long-term response rate of 4-5 years in the 25% range. A limited number of patients in his cohort are beyond five years with minimal disease activity. Of the five “responsive” patients in our trial, one remains with minimal disease activity out 3-4 years from their one-time infusion (participant 6). The other four patients had a full or partial flare of their disease from 18 months to 30 months post single infusion. The response of Participant 1 to retreatment was consistent with prior data from Dr. sun’s group reflecting retreatment is often successful.

There are prior reported trials of MSCs in human lupus; seven used allogeneic derived cells and one used autologous bone marrow-derived cells (31, 40-42). All but two trials were done in China. There is only one “placebo-controlled trial” of MSCs in lupus nephritis patients that were new onset and untreated discussed in the introduction (24, 43). The other two reports of MSCs in lupus were case reports from Europe, one using autologous cells that showed no improvement (40). A more recent paper described compassionate use of UC MSCs in three woman with Class IV lupus nephritis (43). They reported a complete remission in two patients and a partial remission in a third.

The only large placebo-controlled trials reported to date of MSCs in allo and autoimmune diseases, used MSCs for treating Crohns’ fistulas and Graft vs Host disease. The method of derivation and validation of the cells were not described. The MSCs however were late passages and were infused post thawing, both of which are known to impact MSC functionality. These trials showed trends towards efficacy but did not meet their target endpoint. These failures are likely due to the quality of the cells, but led some to postulate MSCs are not effective in immune-mediated diseases (44). There is demonstrated efficacy of MSCs in treating steroid refractory Graft vs Host disease (GvHD) in pediatric patients receiving allogeneic bone marrow. MSCs are approved to treat GvHD in pediatrics in Canada, Japan and New Zealand. MSCs given by direct injection into the local area is approved to treat refractory fistulas in Crohn’s patients in the European Union.

Patients in our trial were on hydroxychloroquine, prednisone and different immunosuppressants. Due to lack of response, two patients had their immunosuppressants discontinued prior to entry into the study. In this limited series, nor in the Sun trials, was there no indication of effects of concomitant medications on responses to MSCs. The impact of concomitant medications on response to MSC therapy is unresolved and must be addressed in future trials.

There are a number of unanswered questions regarding UC MSCs in lupus. The first is how variable are UC MSCs between donors in their efficacy. In this series, we did not see a differential response between the recipients of the two different cord cell lots. In our preclinical studies in lupus-prone mice, we used four different bone marrow donors from controls and three from lupus patients’ (29). The MSCs from controls were more effective in preventing disease progression in the mice than were lupus derived MSCs. The lupus cells had induced indoleamine 2,3-dioxygenase (IDO) expression by gamma interferon and suppressed stimulated T cell proliferation similar to cells from controls. They were, however, not as effective in preventing B cell proliferation (29). Given the prevalence difference in men versus women, one could speculate MSCs derived from males would be more effective than females. We have insufficient numbers to suggest there are differences in MSCs depending on sex of the donor. Defects in lupus MSCs are increasingly reported in vitro (45-47); definitive studies of differences in *in vivo* efficacy of different MSC preparations are lacking. Studies comparing bone marrow-derived versus adipocyte-derived vs UC-derived suggest subtle differences in function, but no definitive data that one source is superior in in vivo trials in humans. We used UC MSCs due to their ready availability, rapid growth characteristics and the ability to treat multiple patients with one cord (>90).

It is clear in humans that following intravenous infusion the majority of the cells are trapped in the lung, but how long they remain viable is unknown (48). It is not known whether the MSCs have to migrate to the affected organ for cell-to-cell interactions for effect or if MSC derived endosomes/cytokines are sufficient (49, 50). In studies in mice infused with human cells, there are reports of a short half-life for the MSCs, while others, including our group, found evidence of MSC survival in target organs for weeks post-infusion (29). The only human study of MSC survival, looked for HLA mismatched MSCs at autopsy of patients having undergone MSC infusion for GvHD (51). MSCs could be detected in different organs weeks after the MSC infusion. Whether cells that are MHC matched or closely matched are preferable to mismatched cells is also unclear, though the “immune privilege” reported for MSCs is time-limited (52). Alloreactivity post-infusion is variable and may or may not enhance the effect.

As controversial as is the topic of efficacy of MSCs, the mechanisms by which they impact disease is also debatable. *In vitro* data indicate that MSCs can suppress the activity of almost every immune cell, while enhancing regulatory B and T cells. A host of mediators are secreted by MSCs including IDO, NO, PGE2, TGFβ, IL10, Factor H and hepatic growth factor (53-55). MSC cell surface molecules such as GARP and FLT3L are postulated to interact with host immune cells impacting proliferation, differentiation and activity (39, 56). Others showed in mice that MSCs are engulfed by resident macrophages inducing a tolerogenic anti-inflammatory phenotype that prolongs the efficacy of MSCs. MSCs are ineffective in mice lacking macrophages (56, 57). At the cellular level, as noted above, MSCs are reported to increase Tregs and Bregs while decreasing Th17 cells, TfH cells and inducing a Th1 to Th2 shift possibly via TGFβ effects (58). Enhancing development of CD1c+ tolerogenic DCs via expression of FLT3L by MSCs was reported, enhancing IFNγ production by CD8+ T cells (56). Although all of the above may contribute, the actual defining mechanisms remain unknown.

Although we did not note any significant changes in the T cell compartment other than in one patient, we did find marked changes in the B cell compartment. The significant effect on DN B cells, aN B cells and SW B cells was not previously reported in published MSC studies. The importance of these findings is supported by the correlation of SLEDAI scores and changes in B cell subsets. The DN B cells and activated naïve cells are increased in active lupus and are believed to be precursors of autoantibody producing cells in lupus (59). Epigenetic analysis of these cells in lupus patients revealed they are primed to respond to TLR ligands, especially TLR7 (35). The lack of notable changes in autoantibodies, despite the B cell shifts, likely reflects the delayed affect any intervention has on autoantibody production, while also supporting the important role of other B cell functions in lupus.

It will be important to determine if the effect of MSCs is a direct effect on B cells or an indirect effect. The impact on B cells seems less likely to be due to T cell effects since we saw evidence of a T cell change in only two patients, while the B cell effect was present in all five responders. Based on the known expression of GARP on MSCs and a prominent role for GARP in tolerance and autoimmunity in mice (39), we assessed GARP-TGFβ levels in participants in this trial. When a GARP bearing cell interacts with another cell that expresses GARP, expression of GARP is increased on both cells (32). This affect may explain the rebound of GARP-TGFβ levels at 24 weeks or may reflect the improvement in disease activity in patients with increased GARP-TGFβ expression.

In summary, our results support the safety of MSCs as a therapy in refractory lupus. Efficacy cannot be determined in this Phase I trial. We, however, considered the response rate encouraging enough to initiate a multi-center double blind placebo-controlled dose trial of UC MSCs compared to standard of care in refractory lupus patients. The B cell and GARP-TGFβ results also support a novel biologic/mechanistic effect of UC-MSCs in patients with lupus. These promising findings are being further studied in the Phase II trial.

## Data Availability

All data produced in the present study are available upon reasonable request to the authors

https://www.MUSC/lupus.edu

## Author contributions

DK was the clinical leader and designer for the trial and performed many of the study visits, CW was involved in mechanistic study design and performing the GARP assays. ZL was involved in the design of the mechanistic studies including the GARP assays. MW was involved in designing the T cell studies and running the assays. CP designed the T cell studies and oversaw their completion. CW designed and oversaw the B cell mechanistic studies, HW was involved in study design and oversaw the production of the MSCs. PN was involved in the study design, and he and EW performed the statistical and methodologic analysis. IS designed and oversaw the B cell mechanistic studies. AR was the lead coordinator and oversaw patient scheduling as well as the design of the trial. GG was the overall study leader and was part of the design of the clinical trial and the mechanistic studies. He also performed patient evaluations.

---

**Supplemental Figure 1.**
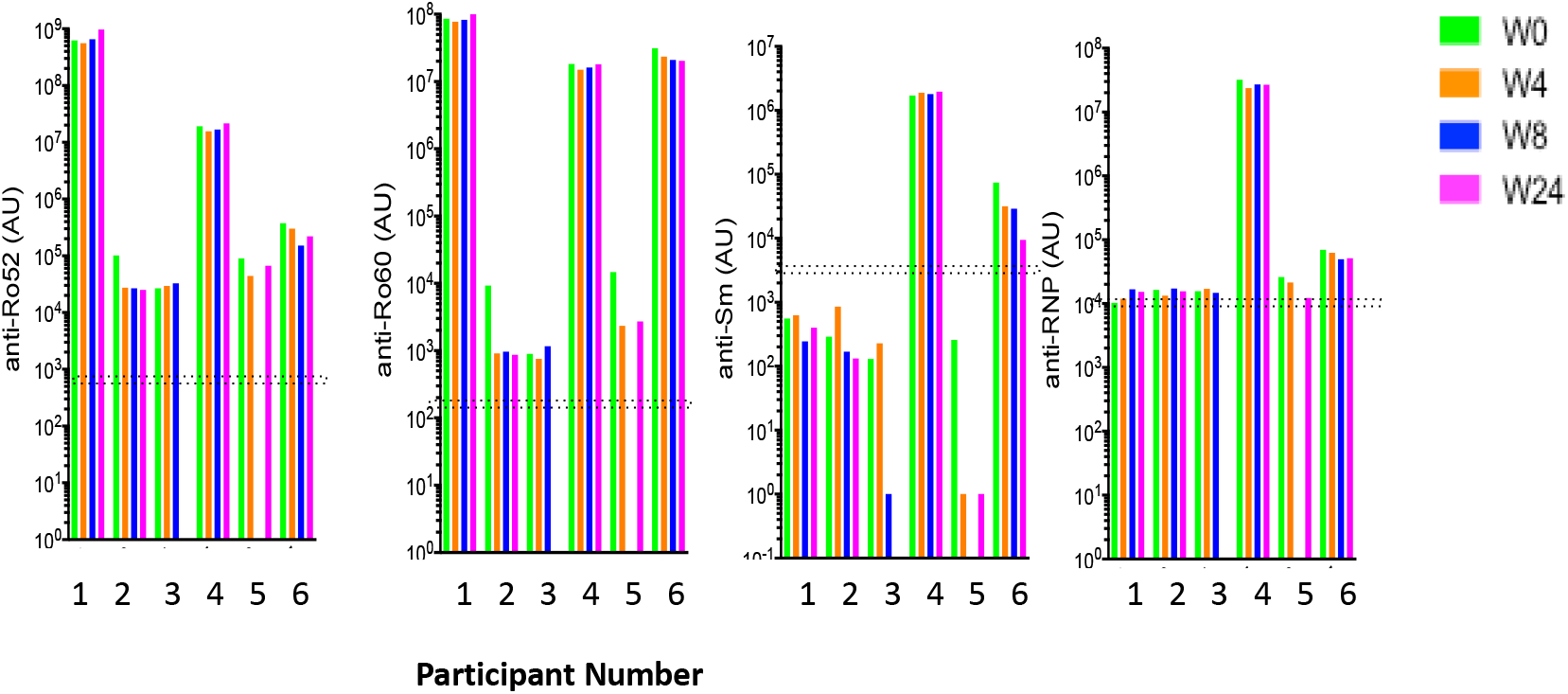
ENA Reactivity over time. Extranuclear nuclear antigen autoantibody assay over time. Anti-Ro52, Ro60, Sm and RNP autoantibodies were determined by the LIPS assay. Reference value for each assay was derived from a group of healthy controls (n=30), and mean+2sd was chosen as the cutoff as indicated by the double dotted line. Week 8 data is missing from some patients due to weather induced missed deliveries of samples.

**Supplemental Figure 2.**
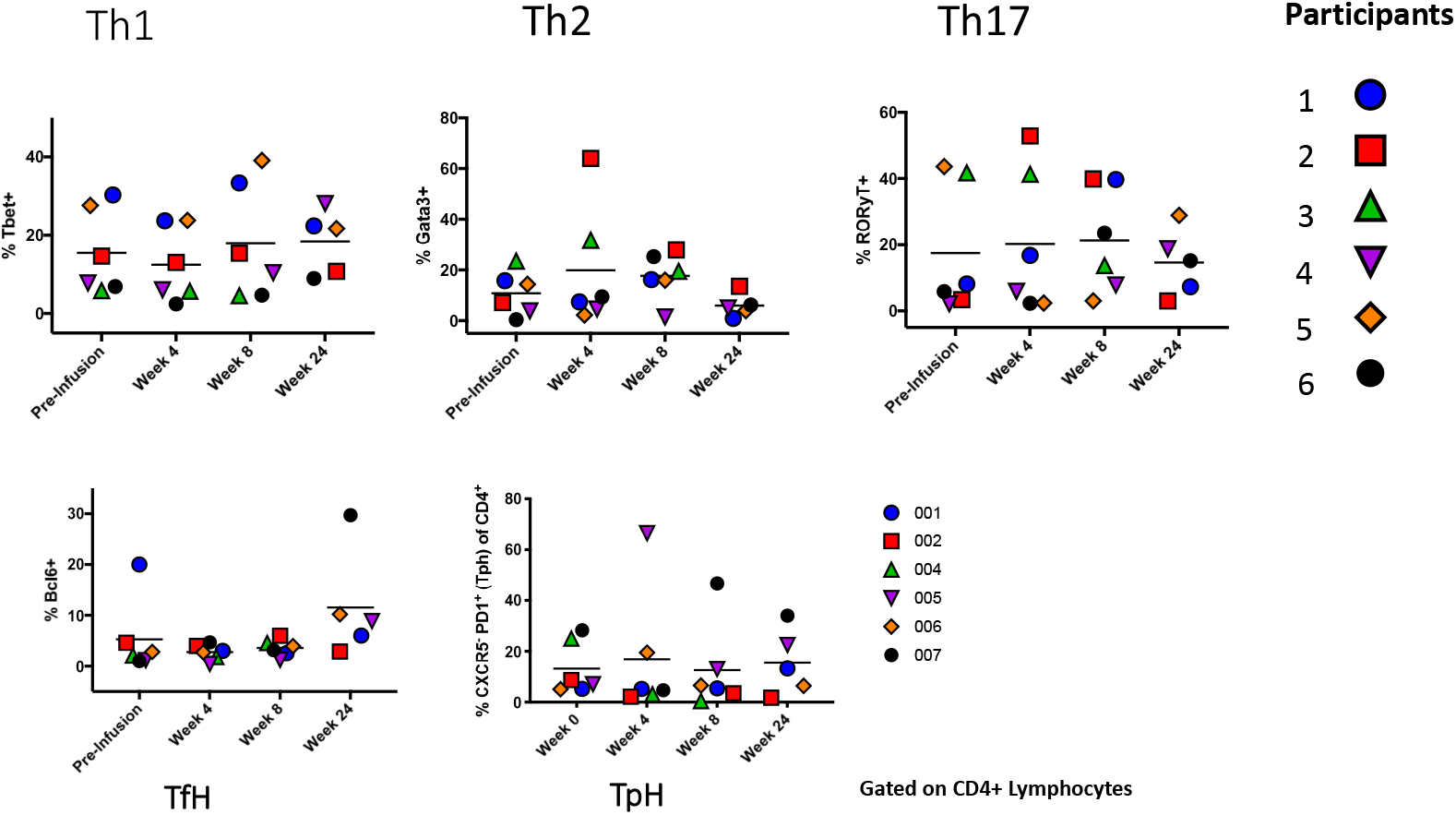
T cell subset changes over time. T cell subset changes over time presented as percent of CD3+ peripheral blood PBMCs. Results using total numbers of T cell subsets revealed similar results to the percentages. There were some individual changes, but overall there was not a significant change in any T cell subset. Results for TfH and TpH subsets are also presented with small overall percent and number and no change overall within the group other than variable changes in individuals that were not consistent.

**Supplemental Table 1:**
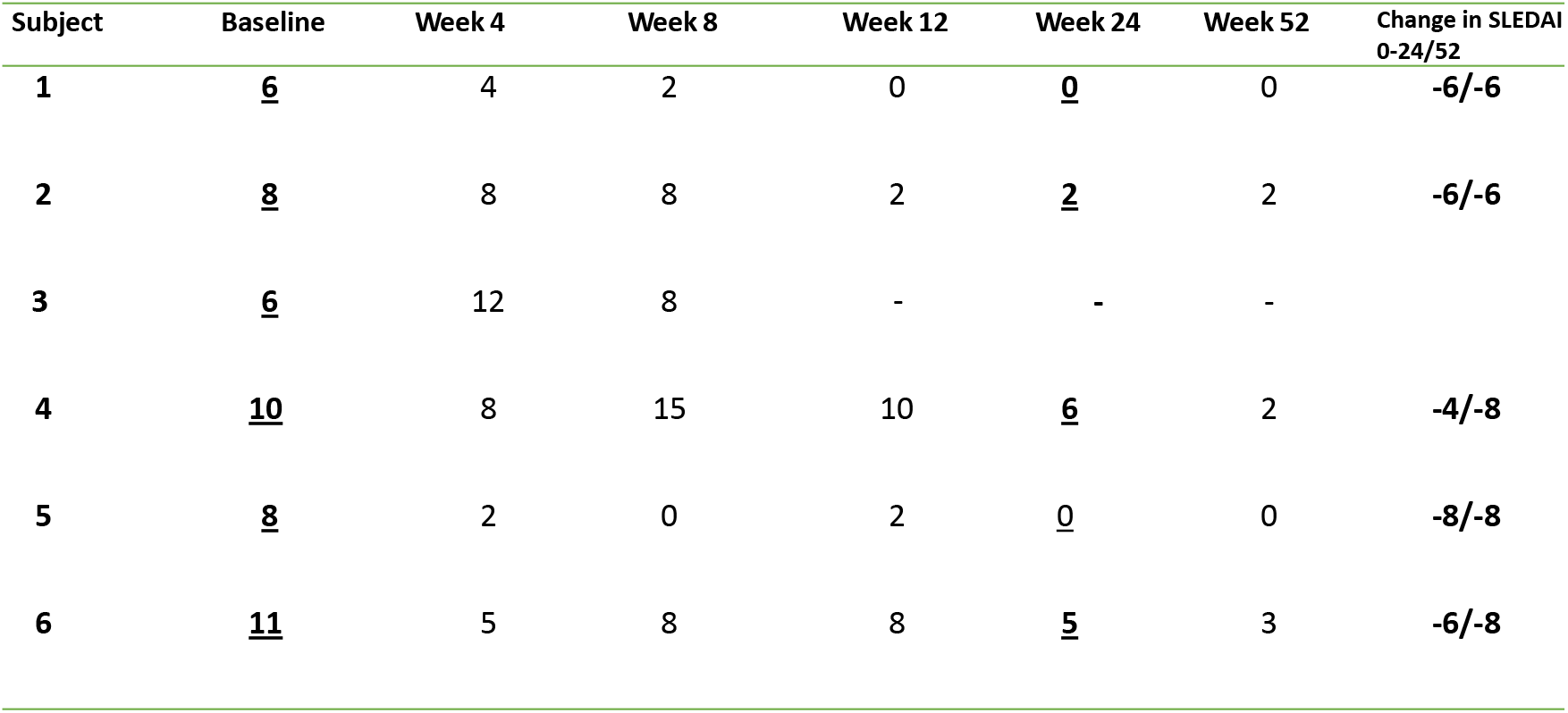
SLEDAI scores at each timepoint. S Table 1 presents the SLEDAI scores of the patients from baseline to week 52. Participant 3 dropped out at week 8 so the data for the following weeks was unattainable. Week 24 was the time when the primary endpoint was determined. The change in SLEDAI score is presented in the final column from baseline to week 24/52.

**Supplemental Table 2:**
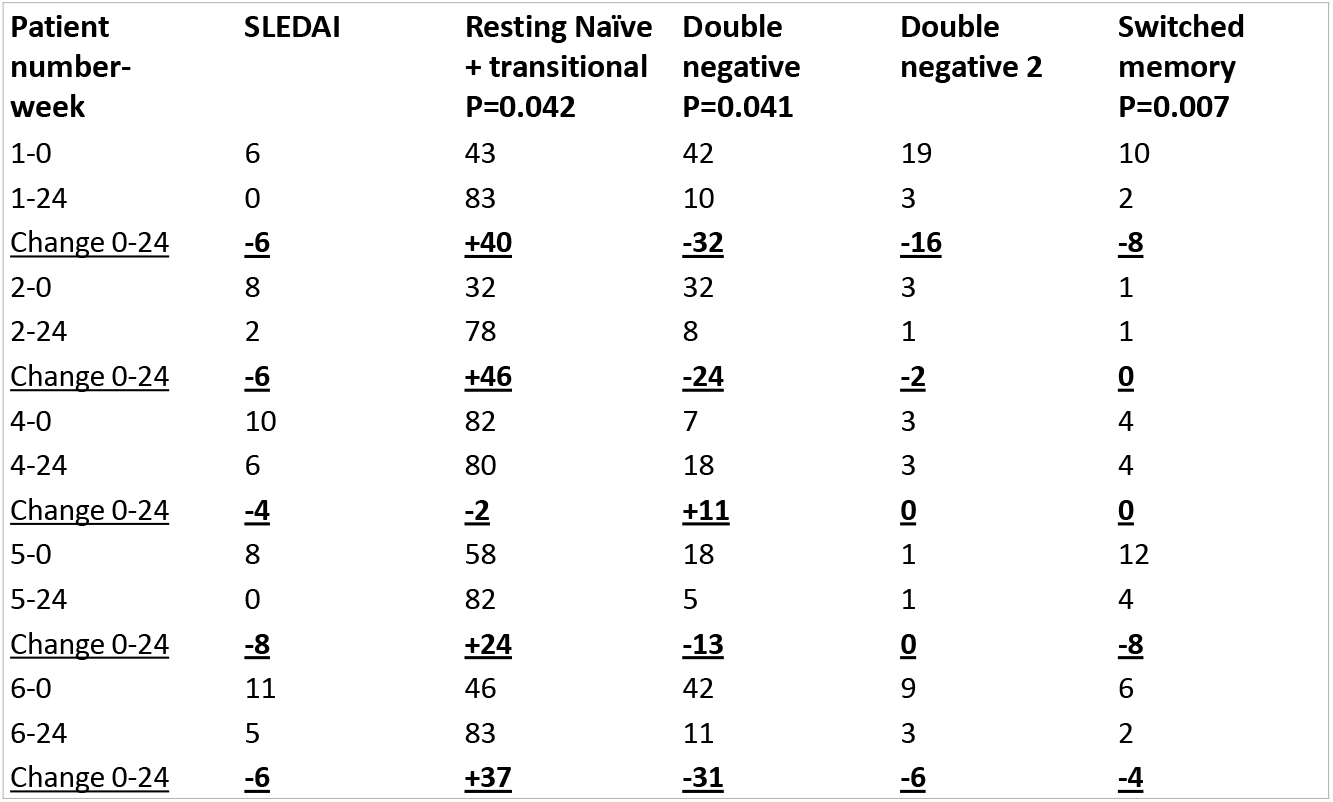
Numerical summary of the B cell subset changes presented graphically in Figure 5. As noted there are significant decreases in double negative B cells and switched memory B cells with a compensatory increase in resting naïve and transitional B cells from baseline to week 24. Data presented are the percent of CD19 B cells. There were no significant changes in overall B cell numbers.

## References

1. Kaul A, Gordon C, Crow MK, Touma Z, Urowitz MB, van Vollenhoven R, et al. Systemic lupus erythematosus. Nat Rev Dis Primers. 2016;2:16039.

2. Ginzler EM, Dooley MA, Aranow C, Kim MY, Buyon J, Merrill JT, et al. Mycophenolate mofetil or intravenous cyclophosphamide for lupus nephritis. N Engl J Med. 2005;353(21):2219–28.

3. Sedhain A, Hada R, Agrawal RK, Bhattarai GR, Baral A. Low dose mycophenolate mofetil versus cyclophosphamide in the induction therapy of lupus nephritis in Nepalese population: a randomized control trial. BMC Nephrol. 2018;19(1):175.

4. Maidhof W, Hilas O. Lupus: an overview of the disease and management options. P T. 2012;37(4):240–9.

5. Miura M, Gronthos S, Zhao M, Lu B, Fisher LW, Robey PG, et al. SHED: stem cells from human exfoliated deciduous teeth. Proc Natl Acad Sci U S A. 2003;100(10):5807–12.

6. Dominici M, Le Blanc K, Mueller I, Slaper-Cortenbach I, Marini F, Krause D, et al. Minimal criteria for defining multipotent mesenchymal stromal cells. The International Society for Cellular Therapy position statement. Cytotherapy. 2006;8(4):315–7.

7. Gronthos S, Mankani M, Brahim J, Robey PG, Shi S. Postnatal human dental pulp stem cells (DPSCs) in vitro and in vivo. Proc Natl Acad Sci U S A. 2000;97(25):13625–30.

8. Lee DE, Ayoub N, Agrawal DK. Mesenchymal stem cells and cutaneous wound healing: novel methods to increase cell delivery and therapeutic efficacy. Stem Cell Res Ther. 2016;7:37.

9. Grada A, Falanga V. Novel Stem Cell Therapies for Applications to Wound Healing and Tissue Repair. Surg Technol Int. 2016;29:29–37.

10. Tse WT, Pendleton JD, Beyer WM, Egalka MC, Guinan EC. Suppression of allogeneic T-cell proliferation by human marrow stromal cells: implications in transplantation. Transplantation. 2003;75(3):389–97.

11. Di Nicola M, Carlo-Stella C, Magni M, Milanesi M, Longoni PD, Matteucci P, et al. Human bone marrow stromal cells suppress T-lymphocyte proliferation induced by cellular or nonspecific mitogenic stimuli. Blood. 2002;99(10):3838–43.

12. Forbes GM, Sturm MJ, Leong RW, Sparrow MP, Segarajasingam D, Cummins AG, et al. A phase 2 study of allogeneic mesenchymal stromal cells for luminal Crohn’s disease refractory to biologic therapy. Clin Gastroenterol Hepatol. 2014;12(1):64–71.

13. Christopeit M, Schendel M, Foll J, Muller LP, Keysser G, Behre G. Marked improvement of severe progressive systemic sclerosis after transplantation of mesenchymal stem cells from an allogeneic haploidentical-related donor mediated by ligation of CD137L. Leukemia. 2008;22(5):1062–4.

14. Wang L, Wang L, Cong X, Liu G, Zhou J, Bai B, et al. Human umbilical cord mesenchymal stem cell therapy for patients with active rheumatoid arthritis: safety and efficacy. Stem Cells Dev. 2013;22(24):3192–202.

15. Wang D, Zhang H, Liang J, Li X, Feng X, Wang H, et al. Allogeneic mesenchymal stem cell transplantation in severe and refractory systemic lupus erythematosus: 4 years of experience. Cell Transplant. 2013;22(12):2267–77.

16. Liang J, Zhang H, Hua B, Wang H, Lu L, Shi S, et al. Allogenic mesenchymal stem cells transplantation in refractory systemic lupus erythematosus: a pilot clinical study. Ann Rheum Dis. 2010;69(8):1423–9.

17. Augello A, Tasso R, Negrini SM, Cancedda R, Pennesi G. Cell therapy using allogeneic bone marrow mesenchymal stem cells prevents tissue damage in collagen-induced arthritis. Arthritis Rheum. 2007;56(4):1175–86.

18. Bonig H, Kuci Z, Kuci S, Bakhtiar S, Basu O, Bug G, et al. Children and Adults with Refractory Acute Graft-versus-Host Disease Respond to Treatment with the Mesenchymal Stromal Cell Preparation “MSC-FFM”-Outcome Report of 92 Patients. Cells. 2019;8(12).

19. Molendijk I, Bonsing BA, Roelofs H, Peeters KC, Wasser MN, Dijkstra G, et al. Allogeneic Bone Marrow-Derived Mesenchymal Stromal Cells Promote Healing of Refractory Perianal Fistulas in Patients With Crohn’s Disease. Gastroenterology. 2015;149(4):918–27 e6.

20. Wang D, Zhang H, Liang J, Wang H, Hua B, Feng X, et al. A Long-Term Follow-Up Study of Allogeneic Mesenchymal Stem/Stromal Cell Transplantation in Patients with Drug-Resistant Systemic Lupus Erythematosus. Stem Cell Reports. 2018;10(3):933–41.

21. Sun L, Wang D, Liang J, Zhang H, Feng X, Wang H, et al. Umbilical cord mesenchymal stem cell transplantation in severe and refractory systemic lupus erythematosus. Arthritis Rheum. 2010;62(8):2467–75.

22. Sun L, Akiyama K, Zhang H, Yamaza T, Hou Y, Zhao S, et al. Mesenchymal stem cell transplantation reverses multiorgan dysfunction in systemic lupus erythematosus mice and humans. Stem Cells. 2009;27(6):1421–32.

23. Gu F, Wang D, Zhang H, Feng X, Gilkeson GS, Shi S, et al. Allogeneic mesenchymal stem cell transplantation for lupus nephritis patients refractory to conventional therapy. Clin Rheumatol. 2014;33(11):1611–9.

24. Deng D, Zhang P, Guo Y, Lim TO. A randomised double-blind, placebo-controlled trial of allogeneic umbilical cord-derived mesenchymal stem cell for lupus nephritis. Ann Rheum Dis. 2017;76(8):1436–9.

25. Musial-Wysocka A, Kot M, Majka M. The Pros and Cons of Mesenchymal Stem Cell-Based Therapies. Cell transplantation. 2019;28(7):801–12.

26. Ankrum JA, Ong JF, Karp JM. Mesenchymal stem cells: immune evasive, not immune privileged. Nat Biotechnol. 2014;32(3):252–60.

27. Abdi R, Fiorina P, Adra CN, Atkinson M, Sayegh MH. Immunomodulation by mesenchymal stem cells: a potential therapeutic strategy for type 1 diabetes. Diabetes. 2008;57(7):1759–67.

28. Wang M, Yuan Q, Xie L. Mesenchymal Stem Cell-Based Immunomodulation: Properties and Clinical Application. Stem Cells Int. 2018;2018:3057624.

29. Collins E, Gu F, Qi M, Molano I, Ruiz P, Sun L, et al. Differential efficacy of human mesenchymal stem cells based on source of origin. J Immunol. 2014;193(9):4381–90.

30. Gao L, Bird AK, Meednu N, Dauenhauer K, Liesveld J, Anolik J, et al. Bone Marrow-Derived Mesenchymal Stem Cells From Patients With Systemic Lupus Erythematosus Have a Senescence-Associated Secretory Phenotype Mediated by a Mitochondrial Antiviral Signaling Protein-Interferon-β Feedback Loop. Arthritis Rheumatol. 2017;69(8):1623–35.

31. Carrion F, Nova E, Ruiz C, Diaz F, Inostroza C, Rojo D, et al. Autologous mesenchymal stem cell treatment increased T regulatory cells with no effect on disease activity in two systemic lupus erythematosus patients. Lupus. 2010;19(3):317–22.

32. Hochberg MC. Updating the American College of Rheumatology revised criteria for the classification of systemic lupus erythematosus. Arthritis Rheum. 1997;40(9):1725.

33. Jolly M, Kosinski M, Garris CP, Oglesby AK. Prospective Validation of the Lupus Impact Tracker: A Patient-Completed Tool for Clinical Practice to Evaluate the Impact of Systemic Lupus Erythematosus. Arthritis Rheumatol. 2016;68(6):1422–31.

34. Fitzmaurice GM, Laird NM, Ware JH. Applied longitudinal analysis. New York: John Wiley & Sons, Inc; 2004.

35. Jenks SA, Cashman KS, Zumaquero E, Marigorta UM, Patel AV, Wang X, et al. Distinct Effector B Cells Induced by Unregulated Toll-like Receptor 7 Contribute to Pathogenic Responses in Systemic Lupus Erythematosus. Immunity. 2020;52(1):203.

36. Wang D, Huang S, Yuan X, Liang J, Xu R, Yao G, et al. The regulation of the Treg/Th17 balance by mesenchymal stem cells in human systemic lupus erythematosus. Cell Mol Immunol. 2015.

37. Tran DQ, Andersson J, Wang R, Ramsey H, Unutmaz D, Shevach EM. GARP (LRRC32) is essential for the surface expression of latent TGF-beta on platelets and activated FOXP3+ regulatory T cells. Proc Natl Acad Sci U S A. 2009;106(32):13445–50.

38. Carrillo-Galvez AB, Cobo M, Cuevas-Ocana S, Gutierrez-Guerrero A, Sanchez-Gilabert A, Bongarzone P, et al. Mesenchymal stromal cells express GARP/LRRC32 on their surface: effects on their biology and immunomodulatory capacity. Stem cells. 2015;33(1):183–95.

39. Wallace CH, Wu BX, Salem M, Ansa-Addo EA, Metelli A, Sun S, et al. B lymphocytes confer immune tolerance via cell surface GARP-TGF-beta complex. JCI Insight. 2018;3(7).

40. Barbado J, Tabera S, Sanchez A, Garcia-Sancho J. Therapeutic potential of allogeneic mesenchymal stromal cells transplantation for lupus nephritis. Lupus. 2018;27(13):2161–5.

41. Woodworth TG, Furst DE. Safety and feasibility of umbilical cord mesenchymal stem cells in treatment-refractory systemic lupus erythematosus nephritis: time for a double-blind placebo-controlled trial to determine efficacy. Arthritis Res Ther. 2014;16(4):113.

42. Zhou T, Li HY, Liao C, Lin W, Lin S. Clinical Efficacy and Safety of Mesenchymal Stem Cells for Systemic Lupus Erythematosus. Stem Cells Int. 2020;2020:6518508.

43. Wen L, Labopin M, Badoglio M, Wang D, Sun L, Farge-Bancel D. Prognostic Factors for Clinical Response in Systemic Lupus Erythematosus Patients Treated by Allogeneic Mesenchymal Stem Cells. Stem Cells Int. 2019;2019:7061408.

44. Galipeau J. The mesenchymal stromal cells dilemma--does a negative phase III trial of random donor mesenchymal stromal cells in steroid-resistant graft-versus-host disease represent a death knell or a bump in the road? Cytotherapy. 2013;15(1):2–8.

45. Nie Y, Lau C, Lie A, Chan G, Mok M. Defective phenotype of mesenchymal stem cells in patients with systemic lupus erythematosus. Lupus. 2010;19(7):850–9.

46. Geng L, Li X, Feng X, Zhang J, Wang D, Chen J, et al. Association of TNF-alpha with impaired migration capacity of mesenchymal stem cells in patients with systemic lupus erythematosus. J Immunol Res. 2014;2014:169082.

47. Zhu Y, Feng X. Genetic contribution to mesenchymal stem cell dysfunction in systemic lupus erythematosus. Stem Cell Res Ther. 2018;9(1):149.

48. Gao J, Dennis JE, Muzic RF, Lundberg M, Caplan AI. The dynamic in vivo distribution of bone marrow-derived mesenchymal stem cells after infusion. Cells Tissues Organs. 2001;169(1):12–20.

49. Karp JM, Leng Teo GS. Mesenchymal stem cell homing: the devil is in the details. Cell Stem Cell. 2009;4(3):206–16.

50. Devine SM, Cobbs C, Jennings M, Bartholomew A, Hoffman R. Mesenchymal stem cells distribute to a wide range of tissues following systemic infusion into nonhuman primates. Blood. 2003;101(8):2999–3001.

51. Le Blanc K, Frassoni F, Ball L, Locatelli F, Roelofs H, Lewis I, et al. Mesenchymal stem cells for treatment of steroid-resistant, severe, acute graft-versus-host disease: a phase II study. Lancet. 2008;371(9624):1579–86.

52. Kean TJ, Lin P, Caplan AI, Dennis JE. MSCs: Delivery Routes and Engraftment, Cell-Targeting Strategies, and Immune Modulation. Stem Cells Int. 2013;2013:732742.

53. Wang D, Feng X, Lu L, Konkel JE, Zhang H, Chen Z, et al. A CD8 T cell/indoleamine 2,3-dioxygenase axis is required for mesenchymal stem cell suppression of human systemic lupus erythematosus. Arthritis Rheumatol. 2014;66(8):2234–45.

54. Meisel R, Zibert A, Laryea M, Gobel U, Daubener W, Dilloo D. Human bone marrow stromal cells inhibit allogeneic T-cell responses by indoleamine 2,3-dioxygenase-mediated tryptophan degradation. Blood. 2004;103(12):4619–21.

55. Singer NG, Caplan AI. Mesenchymal stem cells: mechanisms of inflammation. Annu Rev Pathol. 2011;6:457–78.

56. Yuan X, Qin X, Wang D, Zhang Z, Tang X, Gao X, et al. Mesenchymal stem cell therapy induces FLT3L and CD1c(+) dendritic cells in systemic lupus erythematosus patients. Nat Commun. 2019;10(1):2498.

57. de Witte SFH, Luk F, Sierra Parraga JM, Gargesha M, Merino A, Korevaar SS, et al. Immunomodulation By Therapeutic Mesenchymal Stromal Cells (MSC) Is Triggered Through Phagocytosis of MSC By Monocytic Cells. Stem Cells. 2018;36(4):602–15.

58. Carrion F, Nova E, Luz P, Apablaza F, Figueroa F. Opposing effect of mesenchymal stem cells on Th1 and Th17 cell polarization according to the state of CD4+ T cell activation. Immunol Lett. 2011;135(1-2):10–6.

59. Jenks PJ, Chevalier C, Ecobichon C, Labigne A. Identification of nonessential Helicobacter pylori genes using random mutagenesis and loop amplification. Res Microbiol. 2001;152(8):725–34.

